# Gender and trust in government modify the association between mental health and stringency of social distancing related public health measures to reduce COVID-19: a global online survey

**DOI:** 10.1101/2020.07.16.20155200

**Authors:** Lily O’Hara, Hanan F Abdul Rahim, Zumin Shi

**Author notes:** **Contributors:** HAR and LOH conceptualized the study. All authors conceptualized the analysis plan, and ZS conducted the data analysis. All authors interpreted the results and wrote the manuscript. The corresponding author attests that all listed authors meet authorship criteria and that no others meeting the criteria have been omitted. All authors are guarantors.

## Abstract

**Objectives:** To investigate the associations between stringency of COVID-19 social distancing policies and mental health outcomes, and the moderating effect of trust in government and gender.

**Design and setting:** Cross sectional study involving secondary analysis of publicly available data from a global online COVID-19 survey and the Oxford COVID-19 Government Response Tracker.

**Participants:** 106,497 participants from 58 countries.

**Main outcome measures:** Outcomes were a worries index and a depression index. Predictors were stringency of policies, trust in government, and gender. Multivariable regression was conducted to determine the three-way interaction between the predictor variables for mental health outcomes, adjusting for age, income and education.

**Results:** The median age of participants (56.4% women) was 37 years (interquartile range 29 to 48 years). Women had higher worries and depression scores than men. 45.4% distrusted the government and 43.8% trusted the government to take care of its citizens. Among those who strongly trusted the government, an increase in the stringency of policies was associated with a significant increase in the worries index. Among men who distrusted the government, an increase in policy stringency was associated with an increase in the depression index but not the worries index. In women that strongly distrusted the government, there was an inversed U-shaped association between policy stringency and both the worries and depression indices.

**Conclusion:** As the stringency of public health measures increases, so too do depression and worries. The association is moderated by gender and trust in government. For safe and effective public health measures, governments should develop strategies to increase trust in their actions.

## Introduction

The scale of daily life disruption caused by COVID-19 public health measures is almost unprecedented in modern times, with significant impact on physical, mental and social health and wellbeing.^1^ The ongoing interruption to previous routines, the sense of uncertainty about the future, the financial hardships for individuals, families, and communities, the isolation of physical distancing and working from home, and repeated news consumption about COVID-19 are all factors that contribute to anxiety, stress, and depression in the population.^1^ The public health measures taken by countries affected by COVID-19 have varied significantly, based on the socio-cultural, political, economic, and historical contexts.^2^ Such measures have included a combination of those for people who have tested positive for infection and their contacts, such as quarantine and self-isolation, and those applicable to the public including policies related to behaviours, such as wearing masks and downloading tracking apps, and a range of measures that aim to maximise physical separation between people. Although the purpose is physical distancing, these measures are known as social distancing and are applied across whole populations to reduce the number of times people come into close contact with each other.^3^ Social distancing measures including travel restrictions, orders to shelter in place or stay at home, closing parks, clubs, cafes, restaurants, businesses, schools and universities, cancelling concerts and sporting events, and limiting the number of people allowed in cars, buses, workplaces and supermarkets. It is apparent that some communities have been able to tolerate the long-term application of these public health measures, whilst others have started to voice their objections as life has become incrementally more difficult. Despite some areas of resistance, public health measures have been associated with improved control of the COVID-19 outbreak.^4^ However, the potential population benefits of these public health measures must be weighed very carefully against the negative consequences for mental health and wellbeing.^5^

Research from previous outbreaks, such as Ebola Virus Disease and Zika Virus Disease demonstrated their significant negative impact on mental health and wellbeing, not only in those directly affected by the diseases, but in the general public.^6,7^ Disease outbreaks can trigger new symptoms in those without any previous mental illness and exacerbate symptoms in those with pre-existing illness. Poor mental health outcomes arising from disease outbreaks include fear, depression, anxiety, panic attacks, posttraumatic stress disorder, delirium, psychosis and suicidality.^8^ Some of these outcomes will be triggered by fear of the disease itself and others will result from the public health measures put in place to curb the outbreak. The widespread but relatively understudied impact of disease outbreaks on mental health and wellbeing has been referred to as a ‘second pandemic’^9^ and the ‘forgotten plague’.^7^

In addition to being a health burden in itself, mental health fatigue can be associated with reduced compliance to public health measures. In a study of the lessons learned from Toronto’s mandatory mass quarantine during the severe acute respiratory syndrome (SARS) outbreak in 2003, researchers found that emotional distress tempted some members of the public to consider violating quarantine orders.^10^ Clear information on the protocols and duration of public health measures is one of the key factors for mitigating their negative effects.^11^ People need to understand the measures taken and the rationale for restricting their movements. The negative mental health effects of severe public health measures may be moderated by trust in the government authorities implementing the restrictions. Political trust may change the perception of public health measures from restrictive to protective. Public health and public trust have been described as the defining dyad for the 21^st^ century^12^ and trust in government is regarded as an important determinant of public health outcomes. Declining levels of trust in government are thought to have contributed to declining levels of measles-mumps-rubella (MMR) vaccination and subsequent measles outbreaks in the UK and USA.^13^ On the other hand, high levels of trust in the government may increase compliance to public health measures and mitigate their negative effects. Evidence on this relationship, particularly in times of disease outbreaks, remains surprisingly scarce, however the few studies that have looked at the relationship directly have found such an association. In the early stages of the H1N1 Influenza A pandemic in 2009, trust in the government was positively associated with intentions to adopt protective measures in the Netherlands^14^ and disease avoidance behaviour in the UK.^15^ One of a small number of studies on public health and government trust from lower-income countries showed that Liberians who distrusted the government took fewer precautions against Ebola and were less likely to comply with Ebola control policies.^13^ Cyclical patterns of fear and government distrust were evident in countries most affected by Ebola.^16^

Trust in government is also associated with mental health and wellbeing. In a population-based cross sectional study from southern Sweden, researchers found that lack of trust in the country’s parliament was associated with poor psychological wellbeing.^17^ Low trust in the government after the nuclear disaster in Japan was associated with higher levels of anxiety, particularly for mothers.^18^ In a global survey on COVID-19 attitudes that included over 100,000 participants in 58 countries, trust in the government’s response to COVID-19 was inversely associated with depression and worries about COVID-19.^19^

Successful use of public health measures in a disease pandemic requires reducing, as much as possible the associated negative mental health effects in the population. Although at the time of writing (July 2020), some countries are easing restrictions, others are maintaining strict orders limiting people’s social contact and movement. Even countries that are lifting restrictions must brace themselves for a potential resurgence of cases arising from the premature relaxation of public health measures.^4^ Some areas have reinstated strict measures after an upsurge in cases. Understanding the mechanisms of the negative effects of strict public health measures can help formulate appropriate public health actions before and during a disease outbreak, epidemic or pandemic. The objective of this study was to explore the association between the severity of public health measures and mental health outcomes, including depression and anxiety, during the COVID-19 pandemic, and the moderating role of trust in government in ameliorating those outcomes. As mental health is generally associated with gender, and specifically in the COVID-19 pandemic,^20-25^ we explored associations separately among men and women.

## Methods

### Study period

We undertook secondary analysis of publicly available data collected in a global online survey conducted between 20 March and 7 April 2020. Full description of the survey methodology is available elsewhere.^19^

### Study population

The full dataset included 113,083 participants from 179 countries. Countries with at least 200 participants were included in the current analysis. A total of 960 participants who reported gender other than male or female were excluded from the analysis. A further 2077 participants who completed the survey after April 6 were also excluded. The final analytical sample included 106,497 participants from 58 countries. Countries with the highest number of participants were Brazil, UK and USA (each had more than 11,000) (Supplement Table 1). Of the sample, 100,009 participants from 55 countries could be linked to a country level stringency index.

### Outcome measures

The Patient Health Questionnaire-9 (PHQ-9), a widely used depression scale, was adapted for use in the survey by omitting the item related to suicidal ideation. Respondents indicated how often they had been bothered by specific feelings over the past two weeks. Items included “Little interest in doing things” and “Feeling down, depressed or hopeless” with response options on a four-point scale from ‘Not at all’ to ‘Nearly every day’. Scores for the eight items were averaged and referred to as the depression index (α = .86).

To assess anxieties and worries specific to COVID-19, the survey included five items developed by the researchers, with items such as “I am nervous when I think about current circumstances” and “I am worried about my health” with response options on a five-point scale from ‘Does not apply at all’ to ‘Strongly applies’. Scores for the five items were averaged and referred to as the worries index (α = .58). In the analysis, the depression index and the worries index were converted to z-scores with a mean of 0 and standard deviation of 1.

### Exposure measure

The stringency index was accessed from the Oxford COVID-19 Government Response Tracker (OxCGRT) publicly available data. The OxCGRT project systematically collects information on 17 indicators of public health policy measures that governments have taken to respond to the pandemic. The stringency index includes data on eight containment and closure related public health policies, and one health systems policy on public health information campaigns. Containment and closure public health policies include closure of schools, workplaces and public transport, restrictions on gathering size, internal movement and international travel, cancellations of public events, and stay at home requirements. Each public health policy measure is scored on a scale from 0 to 100 depending on the severity of the policy, and the scores for the nine items are averaged to create the overall stringency index for each country.^26^ Stringency index scores as of 7 April 2020 were included in the analysis.

### Effect modifier

Trust in government was assessed by a single item asking “How much do you trust your country’s government to take care of its citizens?” with response options on a five-point scale from ‘Strongly distrust’ to ‘Strongly trust’.

In the publicly available data, observations from the survey had been re-weighted to make them representative at the country level, based on respondents’ gender, age, income, and education.

### Data analysis

Descriptive analyses were conducted and presented as mean (SD) or percentage. Chi square test and ANOVA analysis were used to compare among groups for categorical and continuous variables, respectively. The factors associated with the trust in government were assessed using multivariable multinominal logistic regression and presented as relative risk ratio (RRR). The factors were mutually adjusted in the multivariable model and presented graphically. The three-way interaction between trust in government, stringency and gender in relation to the worries index and the depression index was examined by adding the product term of the three variables in a multivariable regression model adjusted for age, income (tertile), and education. The results were presented graphically using marginsplot command in Stata. All the analyses were performed by using STATA 16.1 (Stata Corporation, College Station, TX, USA). Statistical significance was considered when p<0.05 (two sided).

## Results

A total of 106,497 participants from 58 countries (56.4% women) were included in the analysis. The median age of the participants was 37 (interquartile range 29 to 48) years (**Table 1**). Two-thirds (67%) of the participants had education levels above 15 years. Women had higher scores on the worries index and depression index than men. Overall, 19.1% participants reported a strong trust in their government, and 26.6% reported strong distrust in the government.

**Table 1.**
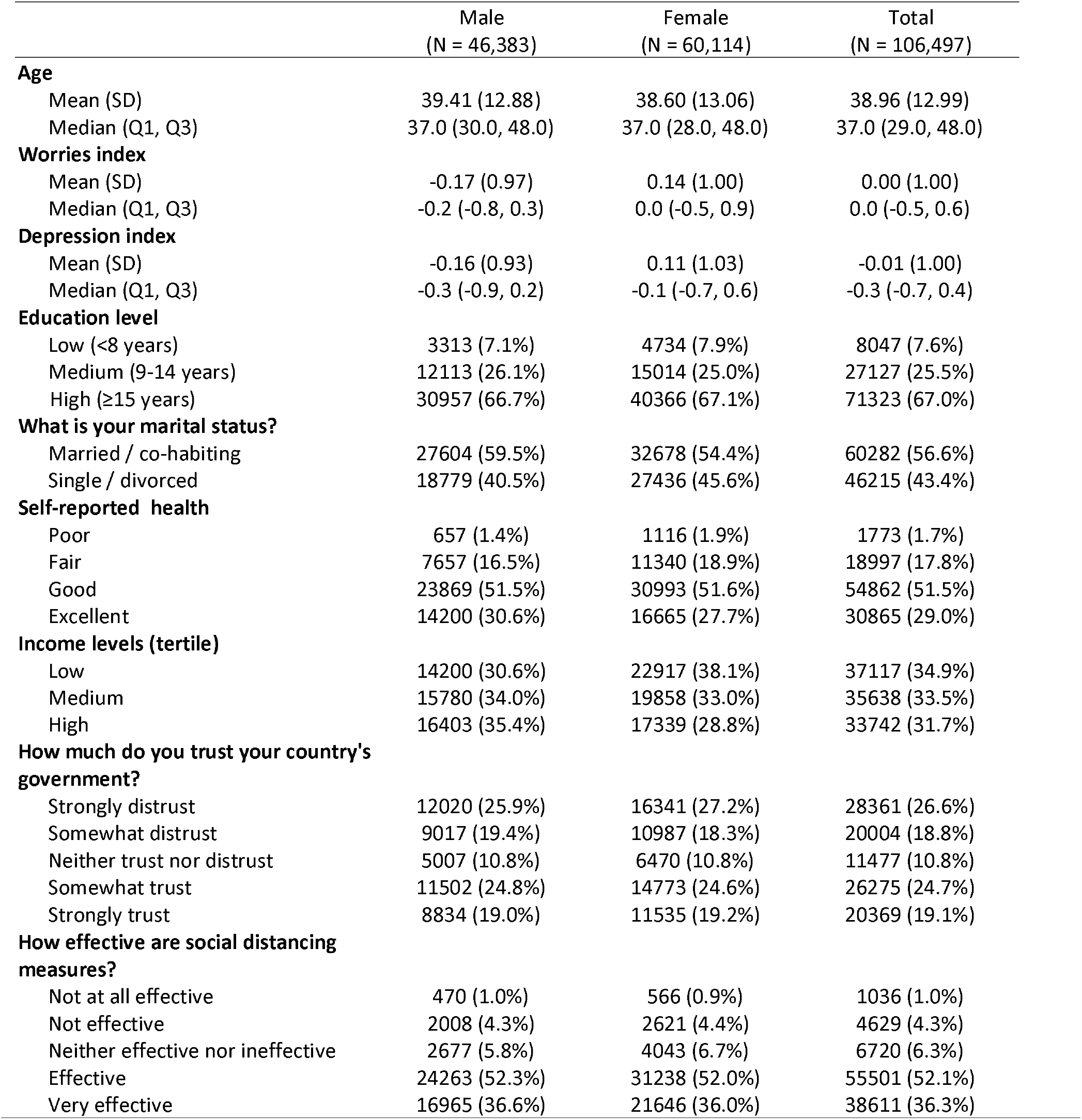
Sample characteristics among participants attending global COVID 19 survey by sex.

**Figure 1** shows the factors associated with trust in the government. Both education and income were positively associated with trust as well as distrust in the government. However, the association between education and distrust in the government was the strongest, whereby those with a high education were more likely to report distrusting their government with a RRR (95%CI) of 1.61(1.48- 1.74). The gender difference in the trust/distrust in the government was small and not statistically significant. The median stringency index was 71.4 (**Supplement Figure 1**).

**Figure 1.**
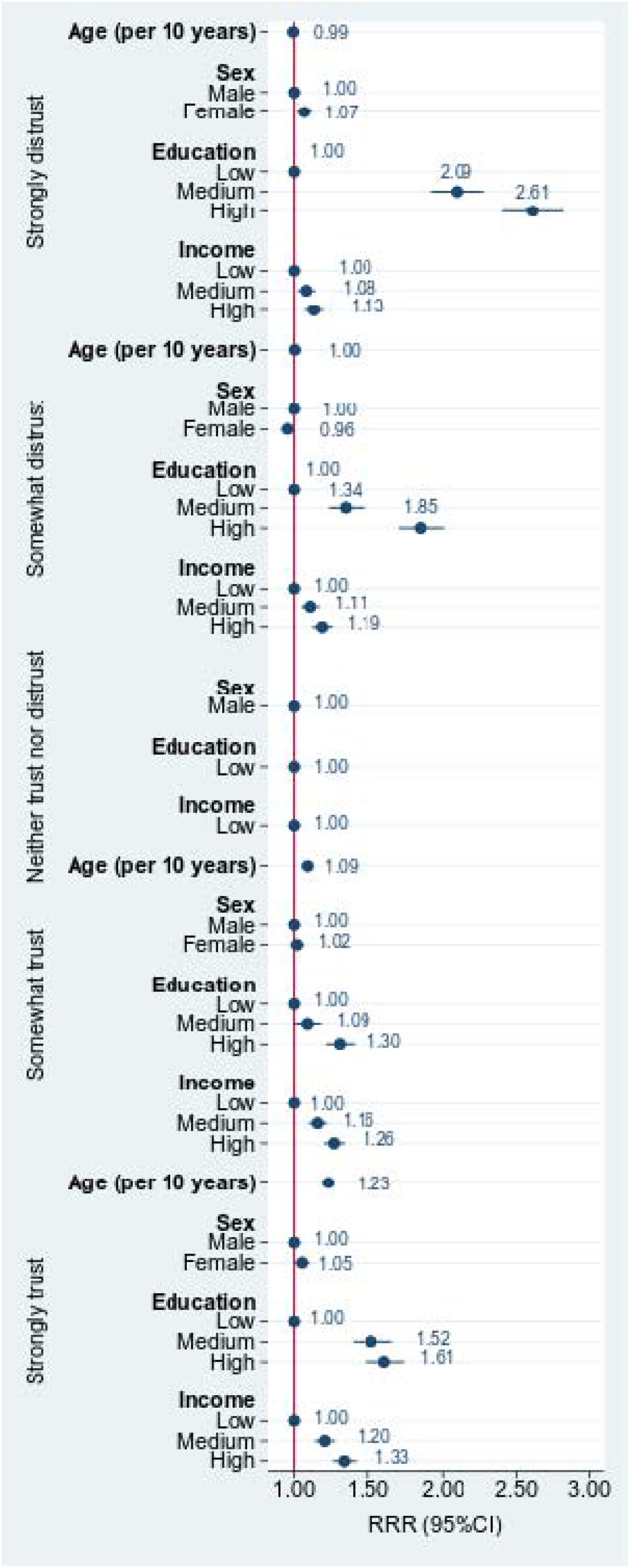
Factors associated with levels of trust in government. Values are RRR (95%CI) from multinominal logistic model adjusting for all the variables in the figure.

In multivariable regression model (**Figure 2**), women and those with a high education had a higher score on the worries index (β 0.30, 95%CI 0.28-0.31) and the depression index (β 0.25, 95%CI 0.24- 0.26). Age and income were inversely associated with both the worries index and depression index.

**Figure 2.**
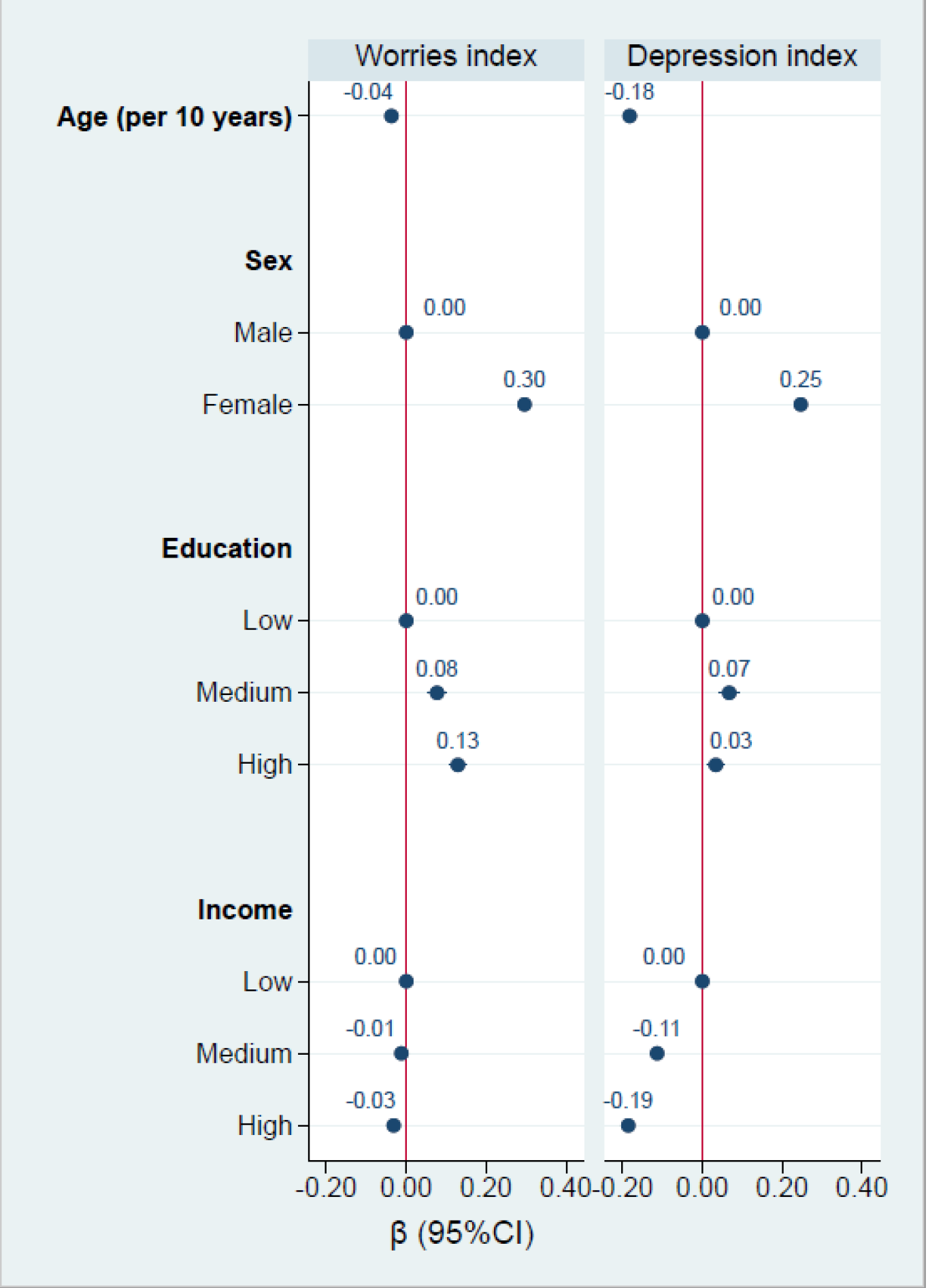
Association between demographic factors and worries index and depression index. All variables in the figure were mutually adjusted.

There was significant a three-way interaction between stringency of public health measures, trust in government and gender in relation to the worries index (**Figure 3**), after adjusting for age, education and income (P<0.001). Overall, both women and men who strongly trusted the government had the lowest worries index, while those who strongly distrusted the government had the highest worries index. With an increase in the stringency index, there was a significant increase in the worries index in women and men who strongly trusted government, but more so in women. Among those who distrusted the government, the increase in the stringency index was not associated with an increase in the worries index in men, while an inversed U-shaped association between the stringency index and worries index was found for women.

**Figure 3.**
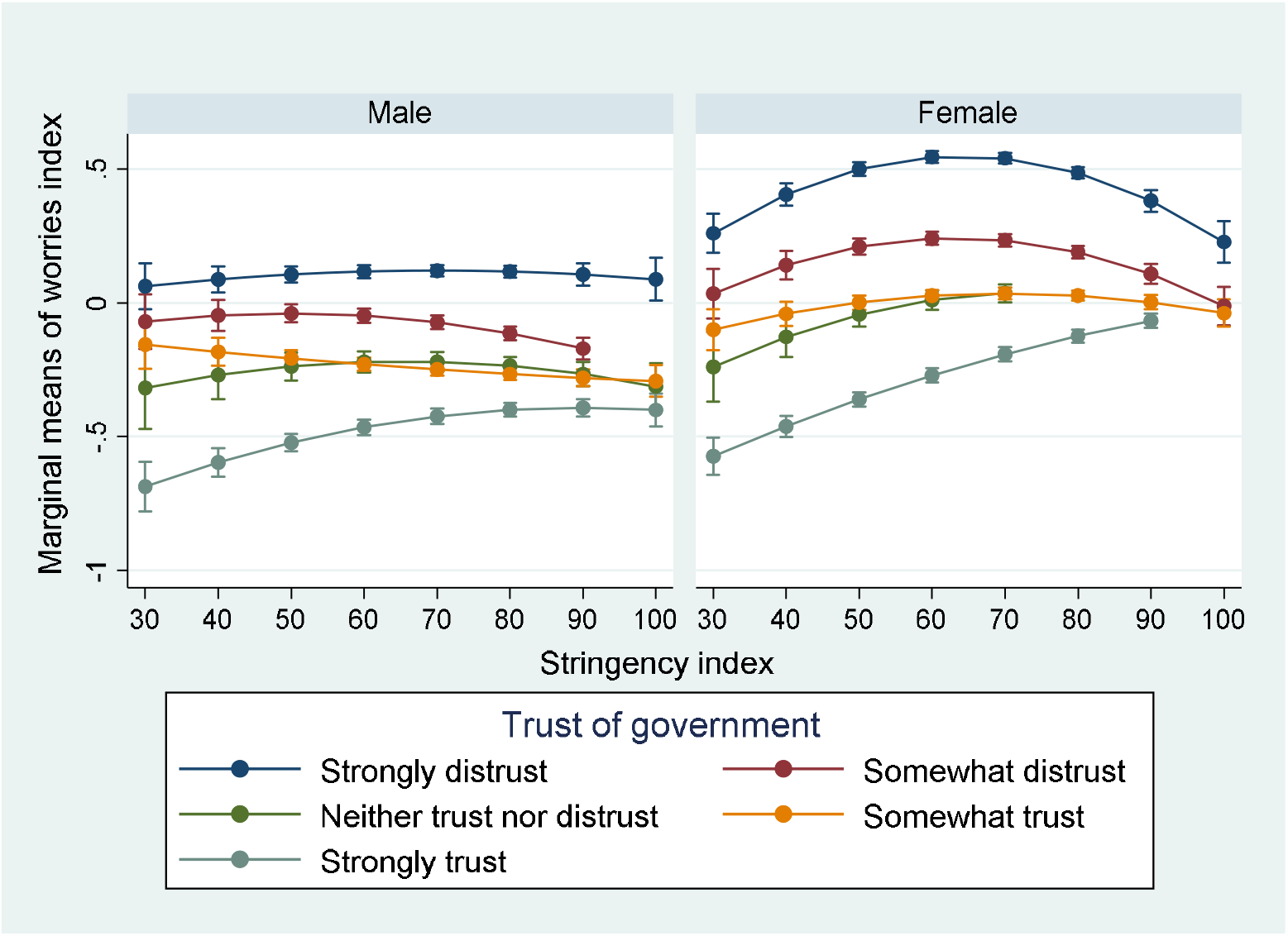
Interaction between trust in government and stringency index in relation to worries index. Model adjusted for age, education and income (country specific tertile). P for gender, trust in government and stringency index interaction <0.001.

There was a significant three-way interaction between stringency of public health measures, trust in the government and gender in relation to the depression index (**Figure 4**) after adjusting for age, education and income (P<0.001). Overall, both women and men who strongly trusted the government had the lowest depression index, while those who strongly distrusted the government had the highest depression index. In men, an increase of the stringency index was associated with an increase in the depression index among those who reported strongly trusting or distrusting their government. In women, a dramatic inversed part U-shaped association between the stringency index and the depression index was observed in those who strongly distrusted their government. Among those who strongly trusted their government, with the increase of the stringency index, there was an increase of the depression index.

**Figure 4.**
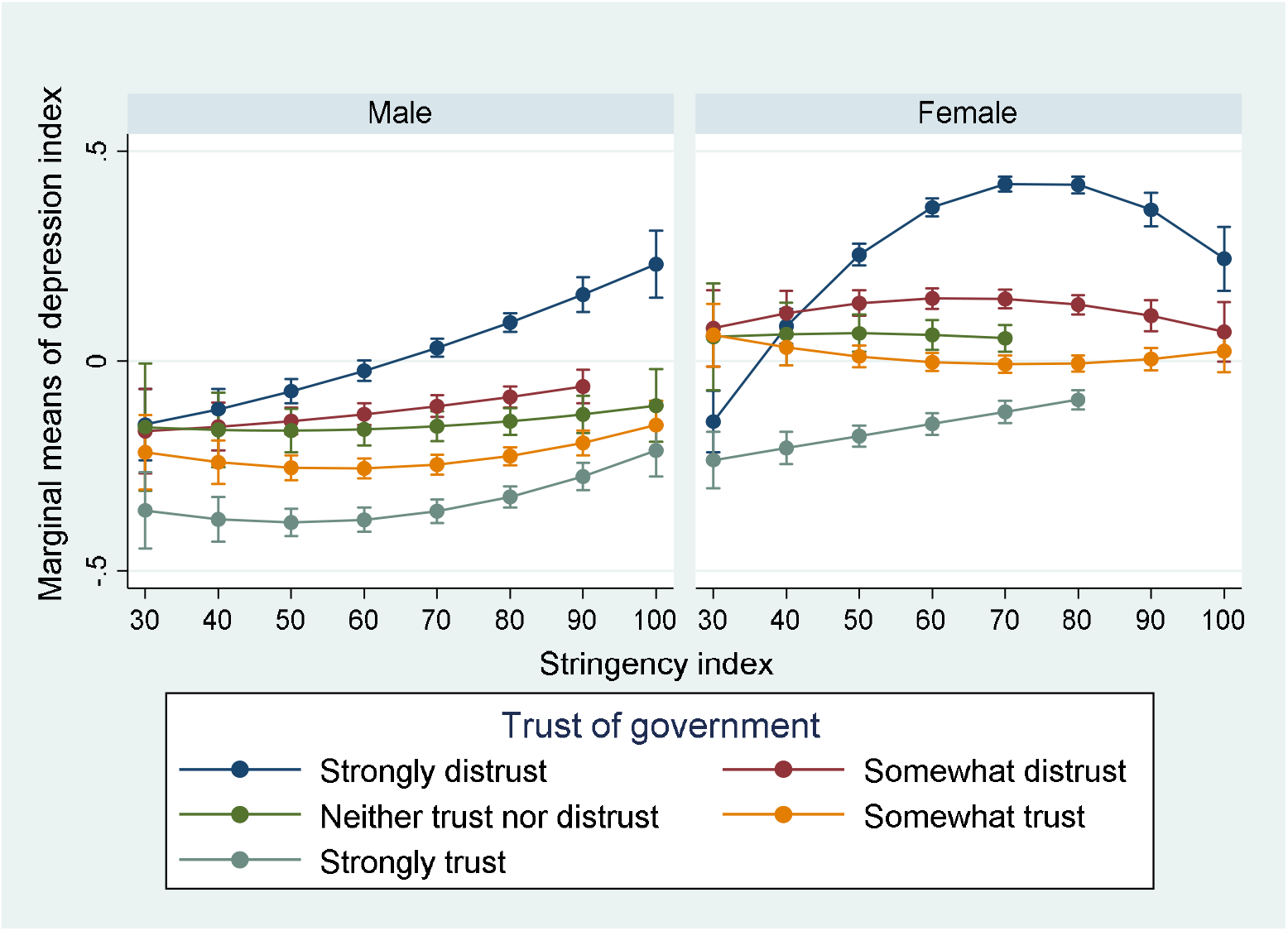
Interaction between trust in government and stringency index in relation to depression index. Model adjusted for age, education and income (country specific tertile). P for gender, trust in government and stringency index interaction <0.001.

There was no three-way interaction between stringency index, trust in government and income in relation to the depression index or worries index (data not shown).

## Discussion

### Principal findings

In this large online survey of participants from 58 countries, there was a high prevalence of strong distrust (26.6%) in government. In contrast, 19.1% of participants strongly trusted their government. Income and education were related to trust in government. Stringency of public health policies was related to mental health indicators for worries and depression. There was a three-way interaction between stringency, gender and trust in government in relation to mental health.

### Comparison with other studies

In this study, women had higher levels of depression and worry than men. This is consistent with findings from other COVID-19 studies from China,^20,22^ Iran,^23^ Italy,^24,25^ and Spain.^21^ For women and men, participants that distrusted the government had the highest levels of depression and worries, and those that trusted the government had the lowest levels of depression and worries. This finding was consistent with studies in Sweden^27^ and Japan^28^ showing positive relationships between levels of trust and mental health.

### Findings of this study

The patterns of relationships between trust and depression and worries were generally true across all levels of stringency of public health measures. However, when analysed by gender, the relationships demonstrated strikingly different patterns for women and men. For women, as the stringency of public health measures increased, both worries and depression indices increased among those who strongly distrust or strongly trust the government. However, for women who strongly distrusted the government there was a tipping point at about the 60 – 70 point stringency mark, after which worries and depression indices decreased. This suggests that for women who strongly distrust the government, once government public health measures become quite strong, there may be a level of security or comfort that this provides that results in better mental health outcomes. For women that distrust, neither trust nor distrust, or trust the government, the stringency of the public health measures makes little difference to worries or depression. For women that strongly trusted the government, worries and depression increased with stringency, but remained the lowest for all categories of women. Overall therefore, for women, more stringent public health measures would be preferred as they result in lower worries and depression than measures that perhaps are regarded as not going far enough.

For men however, a somewhat different picture emerged. As with women, as the stringency of public health measures increased, depression increased for men who strongly distrusted or strongly trusted the government. However, there was no tipping point, and for both categories of men, depression was progressively higher as stringency increased. For men who strongly trusted the government, worries increased with stringency, but remained the lowest for all categories men. However, for those who distrust the government, stricter public health measures mean more imposition and perhaps a perception of more unjustified restrictions.

For men and women who trust or strongly trust the government, more stringent public health measures may result in the perception of the increased seriousness of the pandemic. Because they are more likely to believe that their governments’ actions are reasonable or justified, seeing stricter measures may indicate to them that the pandemic situation is worse. In our analysis, the model is adjusted for income, and therefore any financial distress arising from the public health measures are not likely to be the explanation for these differences.

### Policy implications

These findings present a conundrum for governments that are weighing the costs of public health measures on the health and wellbeing of the population. Once policies exceed the 50 point mark on the stringency index, women benefit from the most stringent policies, yet men do not, particularly men who strongly trust or distrust the government. From both analyses however, it is clear that people with the highest levels of trust had the lowest levels of depression and worries, irrespective of the stringency of the public health measures. In addition to implementing increasingly stringent public health measures to contain the epidemic, governments should concurrently develop and implement strategies to increase trust in their actions. This will maximise the physical and mental health of the community, rather than one at the expense of the other.

As countries roll back some of the closure and containment public health measures, the mental health impact of the measures may begin to reduce. However, if the measures do not remain sufficiently stringent to contain the pandemic, then a second wave of infections may occur^4^ which would undoubtedly increase depression and worries. The World Health Organization has outlined six categories of measures governments should have in place before any rollback of measures begins. The Oxford COVID-19 Government Response Tracker project has conducted an analysis of readiness for countries to rollback restrictive measures using data for four of these categories. As of 1 May 2020, no country had met all four criteria, and yet many were already starting to ease restrictions.^27^

### Strengths and limitations

This study had a number of strengths and limitations. The large number of participants from 58 countries enabled a robust analysis of the relationships between mental health outcomes, trust in government and the stringency of public health measures for women and men. However, the data were from a convenience sample who completed a cross-sectional online survey, and were therefore not representative of the whole population.

### Conclusions

Results from our study suggest that there is a relationship between stringency of public health measures and mental health outcomes. The nature of the association depends on the trust in government and gender as well as the level of the stringency. If the easing of restrictions results in a second wave of infections, governments may need to consider reinstating very stringent public health measures in order to mitigate the mental health impacts on their communities. Building trust in the government to care for its citizens should be considered in order to reduce the risk of mental health problems during the pandemic.

**What is already known on this topic**

- The stringency of public health measures to enhance social distancing has varied according to each country’s rates of COVID-19 and the political, economic and social contexts.
- People’s trust in the government is associated with their willingness to comply with public health measures and with people’s mental health outcomes.
- Gender plays a significant role in mental health outcomes.

**What this study adds**

- Trust in government plays an important and nuanced role in the association between the severity of public health measures to address COVID-19 and mental health outcomes for women and men.
- Governments should focus on developing trust to moderate the negative mental health impacts of strict public health measures to address COVID-19.

## Data Availability

The data used in this study are publicly available

https://osf.io/3sn2k/files/

## Acknowledgments

We gratefully acknowledge the COVID-19 Global Survey team for providing the raw data.

## Patient and public involvement

No patients or members of the public were involved in the design or execution of this study. Since this study used deidentified data, no direct dissemination to research participants is possible.

## Transparency declaration

The corresponding author affirms that the manuscript is an honest, accurate, and transparent account of the study being reported; that no important aspects of the study have been omitted; and that there are no discrepancies from the study as originally planned.

## Ethical approval

Ethical approval was not required for this study as it involved the secondary analysis of de-identified publicly available data.

## Data sharing

The data used in this study are available from https://osf.io/3sn2k/files/

---

**Supplement Table 1.** Distribution of participants by country.

| Country | N |
| --- | --- |
| Brazil | 11,489 |
| United States of America | 11,273 |
| United Kingdom of Great Britain | 11,146 |
| Germany | 9,976 |
| Sweden | 5,780 |
| Switzerland | 4,176 |
| Belarus | 3,643 |
| Russian Federation | 3,380 |
| Mexico | 3,308 |
| Turkey | 2,843 |
| Canada | 2,803 |
| France | 2,703 |
| Spain | 2,314 |
| Peru | 1,971 |
| Colombia | 1,841 |
| Italy | 1,836 |
| Indonesia | 1,596 |
| Ukraine | 1,439 |
| Netherlands | 1,402 |
| Qatar | 1,245 |
| Austria | 1,078 |
| India | 982 |
| Australia | 923 |
| Argentina | 900 |
| Viet Nam | 832 |
| Romania | 803 |
| Finland | 782 |
| Philippines | 734 |
| Ireland | 701 |
| Latvia | 675 |
| Albania | 665 |
| Venezuela, Bolivarian Republic of... | 654 |
| Slovakia | 609 |
| Belgium | 568 |
| Japan | 562 |
| Dominican Republic | 552 |
| Portugal | 552 |
| Chile | 545 |
| South Africa | 541 |
| Malaysia | 528 |
| Denmark | 504 |
| Poland | 476 |
| Singapore | 414 |
| Israel | 406 |
| China | 391 |
| Kenya | 385 |
| Morocco | 372 |
| New Zealand | 352 |
| Greece | 347 |
| Bulgaria | 330 |
| Ecuador | 303 |
| Norway | 300 |
| Thailand | 299 |
| South Korea | 291 |
| Czech Republic | 261 |
| Uruguay | 242 |
| Hungary | 239 |
| Nigeria | 235 |

**Supplement Figure 1.**
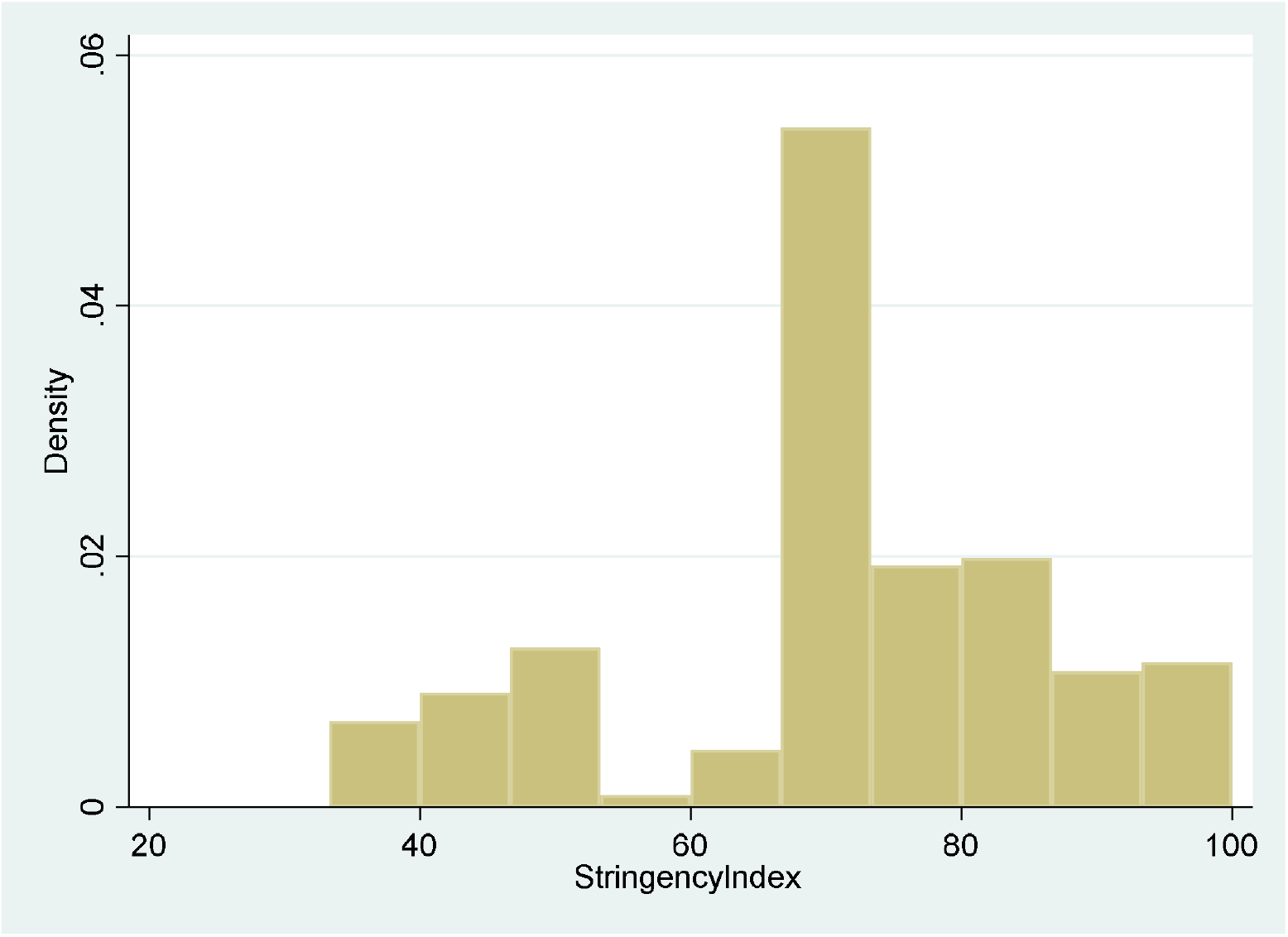
Distribution of stringency index.

